# Deep Fascia Plane Dissection and Mattox Maneuver for Lymphadenectomy in Pancreatic Body–Tail Cancer: SEER-Based Evidence, Anatomical Rationale, and Quantitative Modeling

**DOI:** 10.64898/2026.07.06.26357392

**Authors:** Xinyuan Ye, Yiquan Wang, Wenxian Yang, Jiwang Wu, Jiahe Fang, Given Michael Kihaga, Yihu Zheng

## Abstract

**Introduction:** The optimal examined lymph node (ELN) count after resection for pancreatic body/tail ductal adenocarcinoma (PDAC) is debated. We combined SEER-based survival modelling with a hypothesis-generating theoretical framework to identify ELN thresholds and how surgical approach influences attainability.

**Method:** SEER (2000-2022) resected pancreatic body/tail PDAC cases were analysed (n=5 107). Thresholds were identified by log-rank grid search and segmented Cox changepoint analysis; multivariable Cox and overlap- and inverse-probability-weighted restricted mean survival time (RMST) analyses were performed.

**Results:** Among 5 107 patients, ELN=12 was the optimal cut-point and ELN=21 the changepoint (95% CI 6.0-35.0). Each 5-node increase in ELN was associated with HR 0.964 (95% CI 0.949-0.980); overlap-weighted 60-month RMST gain for ELN>=21 was 2.60 months (95% CI 1.30-3.90). Distribution-derived (model-estimated) ELN>=21 attainment was 16.5% (DP), 40.0% (RAMPS), and 82.7% (n=9, deep retroperitoneal/left-posterior). Theoretical modeling estimated model ELN of 16, 21, and 27 for DP, RAMPS, and Mattox, with Monte Carlo P(ELN>=21) of 5.5%, 34.8%, and 92.9%. Quality-weighted risk reduction was 22.9% for Mattox versus 8.4% for RAMPS.

**Conclusion:** ELN>=12 is a minimum quality benchmark, and approximately 21 a potential higher-yield target. Standard DP carries an accessibility gap for the higher target. Theoretical modeling suggests the Mattox maneuver could bridge this gap, but this remains hypothesis-generating and requires prospective validation.

## Introduction

Pancreatic ductal adenocarcinoma (PDAC) remains one of the most lethal solid malignancies, and tumours of the pancreatic body and tail carry a particularly poor prognosis because they are often diagnosed at an advanced stage [1,2]. Adequate regional lymph node assessment is central to accurate nodal staging, guideline-concordant adjuvant therapy [3], and meaningful survival comparisons across centres and trials [4,5].

Despite this importance, the optimal examined lymph node (ELN) count after distal pancreatectomy remains debated. Several SEER-based and multi-institutional studies have proposed thresholds ranging from 11 to 21 nodes [6–12], and guideline bodies differ: the National Comprehensive Cancer Network (NCCN) recommends examination of at least 12 lymph nodes for adequate staging [13], whereas the International Study Group of Pancreatic Surgery (ISGPS) defines standard lymphadenectomy for left-sided pancreatic cancer by anatomical station groups (stations 10, 11, and 18) rather than by a numeric threshold [14]. For body/tail tumours, anatomical and operative differences mean that thresholds derived from pancreaticoduodenectomy-dominant cohorts may not transfer directly.

Surgical approach may further influence ELN. Radical antegrade modular pancreatosplenectomy (RAMPS) was designed to provide an oncologically en-bloc anterior retroperitoneal dissection, and recent meta-analytic evidence indicates a meaningful ELN gain compared with conventional DP [15–22]. However, the propensity score match analysis by Kwon and colleagues (2024) demonstrated that this ELN gain did not translate into an overall survival benefit, raising the question of whether quantity alone is sufficient or whether the depth of retroperitoneal clearance matters independently [17]. It is important to distinguish artery-first techniques, which optimise vascular control before pancreatic division without altering the dissection plane (Takaori 2016 [27]; Ome 2019 [24]), from a true deep retroperitoneal/left-posterior approach, which enters the deep retroperitoneal plane to access additional nodal stations (no. 8p, 9, 14, 16a2/b1) lying outside the standard anterior dissection plane (Nagai 2020 [23]; Mattox maneuver [39]); direct prospective evidence for left-sided PDAC is scarce and rests on a single small deep-retroperitoneal cohort [23].

The present study therefore combined a SEER-based survival analysis identifying ELN thresholds with a structured evidence synthesis of surgical approach families, and then explored-within an explicitly hypothesis-generating theoretical framework-whether a standardised retroperitoneal approach (Mattox maneuver) could mechanistically explain and bridge the observed accessibility gap. The SEER analysis provides the empirical anchor; the theoretical modeling is presented as a separate, clearly labelled section to avoid conflating observed data with model predictions.

## Methods

### Study design and population

This was a retrospective cohort study using the Surveillance, Epidemiology, and End Results (SEER) 17 registries. Patients aged 18 years or older with microscopically confirmed pancreatic body/tail PDAC (ICD-O-3 codes C25.1-C25.2) who underwent surgical resection between 2000 and 2022 were eligible. Cases with missing or invalid survival duration, examined lymph node count, or positive lymph node count were excluded (n=88); no cases were excluded for positive-to-examined lymph node ratio greater than unity, leaving a final analytic cohort of 5 107 patients. Because SEER contains de-identified publicly available data, institutional review board approval and individual informed consent were not required under the SEER data-use agreement.

### Threshold identification and survival modelling

ELN thresholds were identified by three complementary methods. First, a log-rank grid search across candidate cut-points from 6 to 35 nodes identified the binary threshold with the lowest P value. Second, a segmented Cox proportional hazards model with one changepoint was fitted, and the changepoint location was estimated by maximising the log-likelihood over the candidate range; the 95% confidence interval of the changepoint was obtained by parametric bootstrap with 1 000 replicates. Third, a restricted cubic spline with four knots was fitted within a multivariable Cox model to characterise the nonlinear shape of the ELN-mortality association. Given the exploratory nature of the grid search, P values were not formally adjusted for multiple testing; results were corroborated by the segmented Cox changepoint analysis (Supplementary Table ST01).

Multivariable Cox models adjusted for age (per 10 years), sex, SEER summary stage, primary tumour site (body versus tail), surgical extent, radiotherapy, and chemotherapy. Model discrimination was assessed by the concordance index (C-index) with bootstrap 95% confidence intervals. The proportional hazards assumption was evaluated using Schoenfeld residuals.

### Restricted mean survival time and POS analyses

To quantify absolute survival differences, restricted mean survival time (RMST) was estimated at 12, 36, and 60 months for ELN thresholds of 12, 14, and 21 nodes. Both overlap weighting (OW) and inverse-probability-weighting (IPTW) were used to balance baseline covariates between high- and low-ELN groups. Delta RMST and bootstrap 95% confidence intervals are reported.

For positive-node-count (POS) analysis, a Cox model incorporating a step term (POS=0 versus POS>=1) and a continuous spline term was used to distinguish the step change at the node-positive threshold from the graded risk across higher POS values. Delta RMST was also computed for POS tiers relative to POS=0.

### Surgical approach evidence synthesis

A structured evidence synthesis of 17 studies [12,16–19,22–32,36] contextualised attainability of the SEER-derived ELN thresholds by approach. Studies were grouped into three families: conventional DP, RAMPS (including the antegrade and retrograde variants), and deep retroperitoneal/left-posterior approaches (including the left-posterior route and the Mattox maneuver). Two artery-first technical reports (Takaori 2016 [27]; Ome 2019 [24]) were catalogued for mechanistic context only and contribute no ELN-based quantitative data to the pooled estimates. Pooled ELN distributions and the proportion of patients achieving ELN>=21 were computed for each family. Distribution-derived (model-estimated) and model-based (Monte Carlo) attainment rates are reported. Study-level means and SDs were harmonized from reported medians and ranges: means by the Hozo et al. formula (5), (a+2m+b)/4 [40], and SDs by the Wan et al. sample-size-dependent estimator [41]; attainment probabilities were derived as right-tail areas of the estimated normal distribution with a 0.5 continuity correction; these are model-estimated values, not observed proportions. Minagawa 2022 and Nagai 2020 served as evidence inputs and theoretical modeling anchors; their dual use constitutes partial overlap, not independent validation. (Supplementary Table ST03 catalogues all 17 studies; Supplementary Table ST09 reports the 14 estimable arms.)

### Theoretical modeling framework

A six-model theoretical framework was constructed to estimate whether a standardised retroperitoneal approach could bridge the accessibility gap. The framework comprises: (1) anatomical accessibility; (2) counterfactual falsification (Delta-S removal); (3) dose-response modeling (RRR); (4) N+ detection probability; (5) Monte Carlo simulation (10 000 runs); and (6) HR decomposition into quantity and quality components. All Mattox-related numerical estimates are theoretical predictions, not clinical observations, and are presented as hypothesis-generating only (Supplementary Table ST08).

Internal validation comprised one-at-a-time parameter perturbation, parametric bootstrap (500 replicates, 1 000 inner simulations), and cross-validation against external evidence (Minagawa median ELN [12], Portelli meta-analytic ELN difference [33], Nagai left-posterior ELN [23]). Analyses used Python 3.10 (lifelines, pandas, numpy, scikit-learn). Two-sided P<0.05 was considered significant.

## Results

### Cohort characteristics and ELN distribution

The final cohort comprised 5 107 patients with resected pancreatic body/tail PDAC (Table 1). There were 3 630 deaths (71.1%) during the study period. Median follow-up was 70.9 months (IQR 32.7-123.9), calculated by the reverse Kaplan-Meier method. The median ELN was 14 nodes (interquartile range 8-21), and the distribution was right-skewed. Patients with ELN>=12 were slightly younger and more likely to be female than those with ELN<12, with small standardised mean differences (SMD<0.10) for most baseline covariates.

**Table 1.** Baseline characteristics of the SEER cohort by ELN<12 versus ELN>=12 stratum (n=5 107).

| Variable /<br>Category<br>(ref: ELN<12) | ELN<12 (n=1<br>920)<br>(n=1 920) | ELN>=12 (n=3<br>187)<br>(n=3 187) | P value<br>(<12 vs >=12) | SMD<br>(<12 vs >=12) |
| --- | --- | --- | --- | --- |
| Age |  |  | 0.0025 | 0.081 |
| (24.999, 60.0] | 651 (33.9%) | 1157 (36.3%) |  |  |
| (60.0, 65.0] | 344 (17.9%) | 636 (20.0%) |  |  |
| (65.0, 75.0] | 667 (34.7%) | 1025 (32.2%) |  |  |
| (75.0, 90.0] | 258 (13.4%) | 369 (11.6%) |  |  |
| Sex |  |  | 0.0021 | 0.073 |
| Female | 938 (48.9%) | 1700 (53.3%) |  |  |
| Male | 982 (51.1%) | 1487 (46.7%) |  |  |
| Unknown | 0 (0.0%) | 0 (0.0%) |  |  |
| Race |  |  | 0.5737 | 0.023 |
| White | 1540 (80.2%) | 2570 (80.6%) |  |  |
| Black | 185 (9.6%) | 328 (10.3%) |  |  |
| Asian/Pacific<br>Islander | 180 (9.4%) | 269 (8.4%) |  |  |
| AI/AN | 13 (0.7%) | 15 (0.5%) |  |  |
| Unknown | 2 (0.1%) | 5 (0.2%) |  |  |
| Grade |  |  | 0.0016 | 0.058 |
| Grade I | 182 (9.5%) | 254 (8.0%) |  |  |
| Grade II | 930 (48.4%) | 1573 (49.4%) |  |  |
| Grade III | 550 (28.6%) | 846 (26.5%) |  |  |
| Grade IV | 20 (1.0%) | 18 (0.6%) |  |  |
| Unknown | 238 (12.4%) | 496 (15.6%) |  |  |
| Tumour size |  |  | 0.0018 | 0.073 |
| <=2 cm | 253 (13.2%) | 433 (13.6%) |  |  |
| >2-4 cm | 802 (41.8%) | 1489 (46.7%) |  |  |
| >4 cm | 822 (42.8%) | 1206 (37.8%) |  |  |
| Unknown | 43 (2.2%) | 59 (1.9%) |  |  |
| Stage |  |  | <0.001 | 0.240 |
| Localized | 270 (14.1%) | 262 (8.2%) |  |  |
| Regional | 855 (44.5%) | 1229 (38.6%) |  |  |
| Distant | 305 (15.9%) | 311 (9.8%) |  |  |
| Unknown | 490 (25.5%) | 1385 (43.5%) |  |  |
| Primary site |  |  | <0.001 | 0.228 |
| Body | 651 (33.9%) | 1513 (47.5%) |  |  |
| Tail | 1269 (66.1%) | 1674 (52.5%) |  |  |
| Other | 0 (0.0%) | 0 (0.0%) |  |  |
| Positive nodes (POS) |  |  | <0.001 | 0.247 |
| POS=0 | 1082 (56.4%) | 1375 (43.1%) |  |  |
| POS=1-2 | 613 (31.9%) | 975 (30.6%) |  |  |
| POS=3-4 | 169 (8.8%) | 404 (12.7%) |  |  |
| POS>=5 | 56 (2.9%) | 433 (13.6%) |  |  |
| Surgery extent |  |  | 0.1519 | 0.033 |
| Local | 1173 (61.1%) | 1912 (60.0%) |  |  |
| Combined | 639 (33.3%) | 1099 (34.5%) |  |  |
| Extended | 101 (5.3%) | 173 (5.4%) |  |  |
| Other | 7 (0.4%) | 3 (0.1%) |  |  |
| Radiation |  |  | 0.001 | 0.095 |
| Yes | 512 (26.7%) | 719 (22.6%) |  |  |
| No/Unknown | 1408 (73.3%) | 2468 (77.4%) |  |  |
| Chemotherapy |  |  | <0.001 | 0.216 |
| Yes | 1301 (67.8%) | 2465 (77.3%) |  |  |
| No/Unknown | 619 (32.2%) | 722 (22.7%) |  |  |
| Year |  |  | <0.001 | 0.261 |
| 2000-2004 | 246 (12.8%) | 120 (3.8%) |  |  |
| 2005-2009 | 420 (21.9%) | 355 (11.1%) |  |  |
| 2010-2014 | 464 (24.2%) | 697 (21.9%) |  |  |
| 2015-2019 | 503 (26.2%) | 1131 (35.5%) |  |  |
| 2020-2022 | 287 (14.9%) | 884 (27.7%) |  |  |

Abbreviations: ELN, examined lymph nodes; SEER, Surveillance, Epidemiology, and End Results; SMD, standardized mean difference. Categorical variables are presented as n (%). P values were calculated using the chi-square test and are rounded to four decimals; values below 0.001 are reported as <0.001. SMDs quantify between-group imbalance.
Note: Standardized mean differences (SMD) exceeding 0.10 for stage, primary tumour site, chemotherapy, positive-node count, and year reflect the expected association between higher lymph node yield and more advanced disease or more recent treatment era. In the restricted mean survival time analyses (Table 2), overlap and inverse-probability weighting reduced all post-weighting SMDs to $\leq 0.01$ .

### ELN thresholds and nonlinear mortality association

The log-rank grid search identified ELN=12 as the optimal binary cut-point within the candidate range of 6-35 nodes (Figure 1A). Segmented Cox analysis identified a single changepoint at ELN=21 (bootstrap 95% CI 6.0-35.0; Figure 1B). The restricted cubic spline showed a steeper inverse association below 21 nodes with an attenuated slope above this threshold (Figure 1C), consistent with a dual-threshold interpretation. In the multivariable Cox model, each 5-node increase in ELN was associated with a 3.6% lower hazard of death (HR 0.964, 95% CI 0.949-0.980; Figure 1D), with C-index 0.659 (95% CI 0.650-0.669).

**Figure 1.**
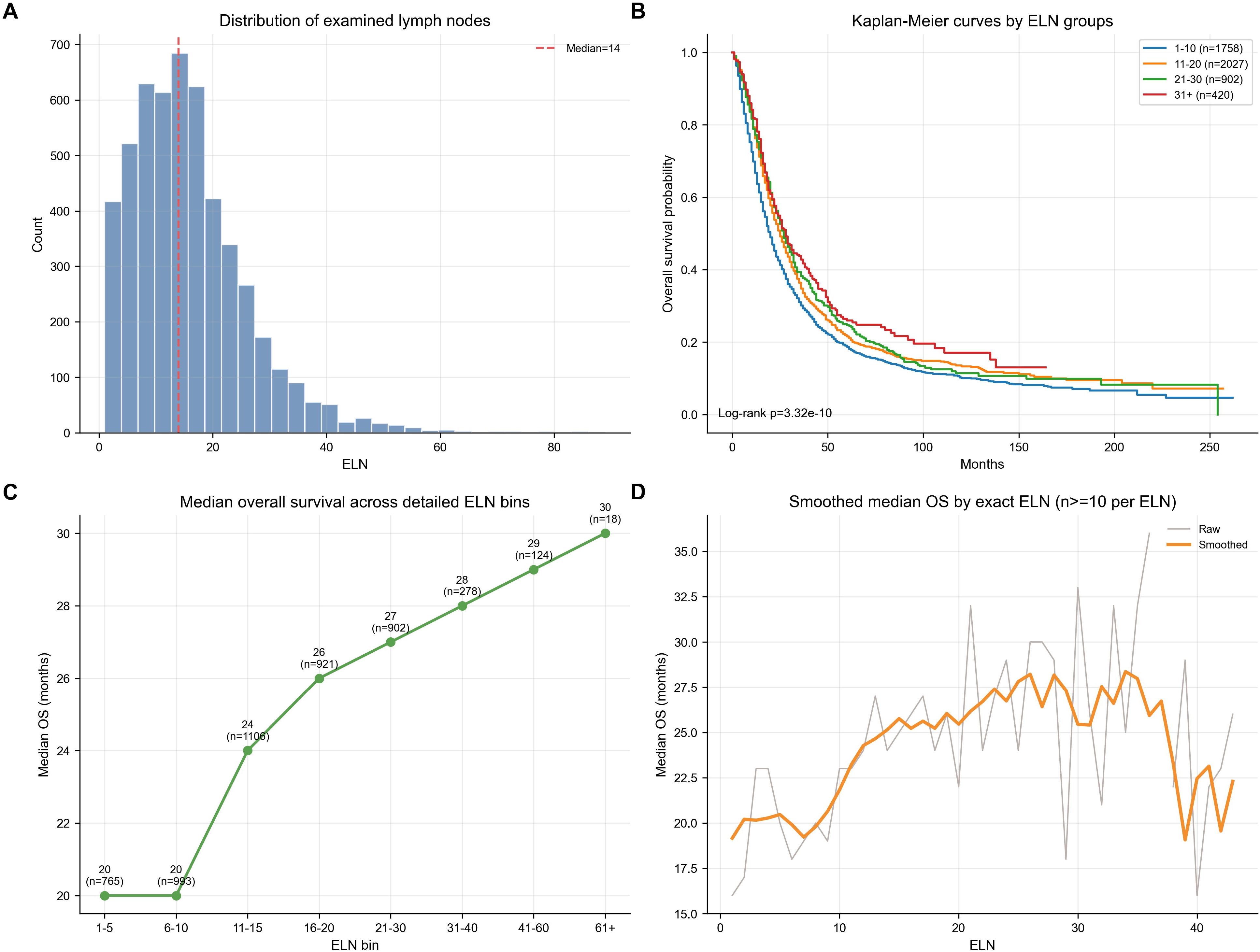
ELN threshold identification by multiple methods (SEER cohort, n=5 107). Panel A: log-rank grid search P values across candidate cut-points, identifying ELN=12 as the optimal binary threshold. Panel B: segmented Cox changepoint analysis, identifying ELN=21 as the changepoint (bootstrap 95% CI 6.0-35.0). Panel C: restricted cubic spline showing the nonlinear ELN-mortality association, with steeper slope below 21 and attenuation above. Panel D: multivariable Cox model adjusted hazard ratio per 5-node increase in ELN (HR 0.964, 95% CI 0.949-0.980), with model C-index 0.659 (95% CI 0.650-0.669). Alt text: ELN threshold identification in SEER cohort. Panel A: log-rank grid search. Panel B: segmented Cox changepoint. Panel C: spline HR curve. Panel D: RMST curves.

### RMST and survival differences

At 60 months, overlap-weighted RMST gains were 2.59 months for ELN>=12 (95% CI 1.43-3.76; P=0.001), 2.31 months for ELN>=14 (95% CI 1.02-3.52), and 2.60 months for ELN>=21 (95% CI 1.30-3.90; Table 2). Corresponding IPTW estimates were directionally consistent, with 2.91 months for ELN>=21 (95% CI 1.61-4.42). The ELN>=21 60-month delta-RMST of 2.60 months is used as an empirical anchor in the Discussion; it is a SEER-observed association, not an expected treatment effect.

**Table 2.**
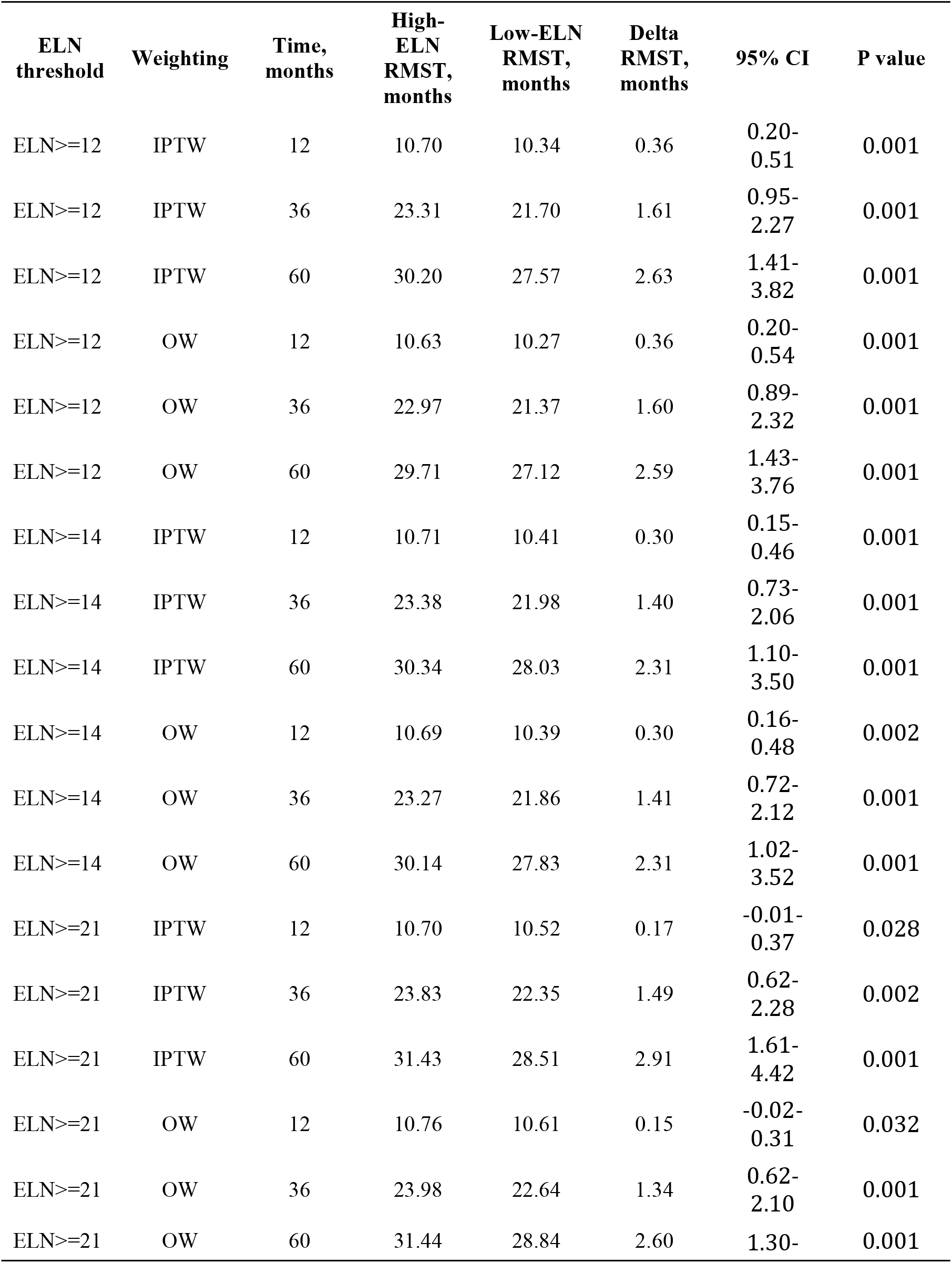
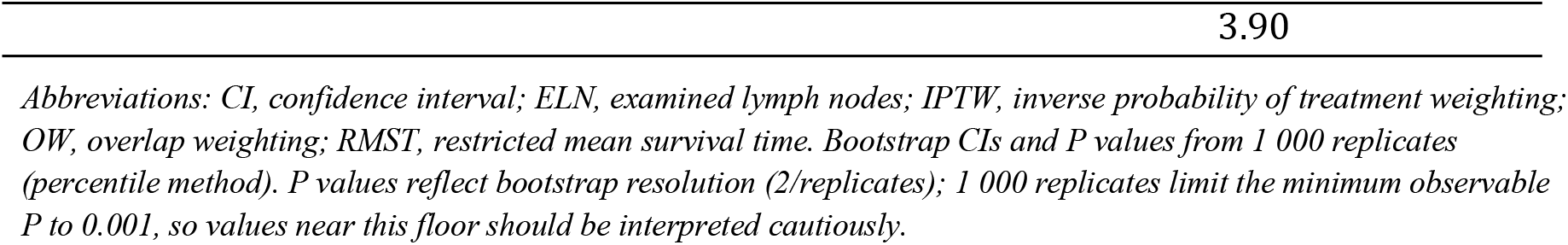
Overlap-weighted and inverse-probability-weighted restricted mean survival time (RMST) differences at 12, 36, and 60 months for ELN thresholds of 12, 14, and 21 nodes. Bootstrap 95% confidence intervals and P values were derived from 1 000 bootstrap replicates.

### Positive lymph node count and risk step

Among all 5 107 patients with complete POS information, a clear risk step was observed from POS=0 to POS>=1 (HR 1.403, 95% CI 1.314-1.499; supplementary figures). Beyond the step, additional POS was associated with graded risk: POS 1-2 versus POS=0 yielded a 60-month RMST loss of 5.81 months, and POS>=5 yielded a loss of 12.22 months. The biological plausibility of an ELN threshold is therefore supported by the observation that more extensive retrieval increases the probability of correctly identifying node-positive disease.

### Surgical approach and model-estimated threshold attainment

The structured evidence synthesis included 17 studies [12,16–19,22–32,36] (Figure 2; Supplementary Table ST03, ST04, ST09). Pooled distribution-derived (model-estimated) attainment rates for ELN>=21 were 16.5% for conventional DP, 40.0% for RAMPS, and 82.7% for deep retroperitoneal/left-posterior approaches (a single cohort, n=9) (Figure S5; Supplementary Table ST07). The deep retroperitoneal/left-posterior family therefore approaches the SEER-derived higher-yield threshold in model-estimated attainment, whereas the conventional DP family remains below it (Supplementary Table ST06). The Kwon 2024 propensity score match analysis reported ELN of 15.0 for RAMPS versus 10.0 for cDPS (P=0.001), R0 rates of 94.9% versus 93.9% (P=0.756), and 5-year overall survival of 45.2% versus 44.4% (P=0.853), indicating that the ELN gain achieved by an anterior approach modification alone (RAMPS) did not translate into a survival benefit.

**Figure 2.**
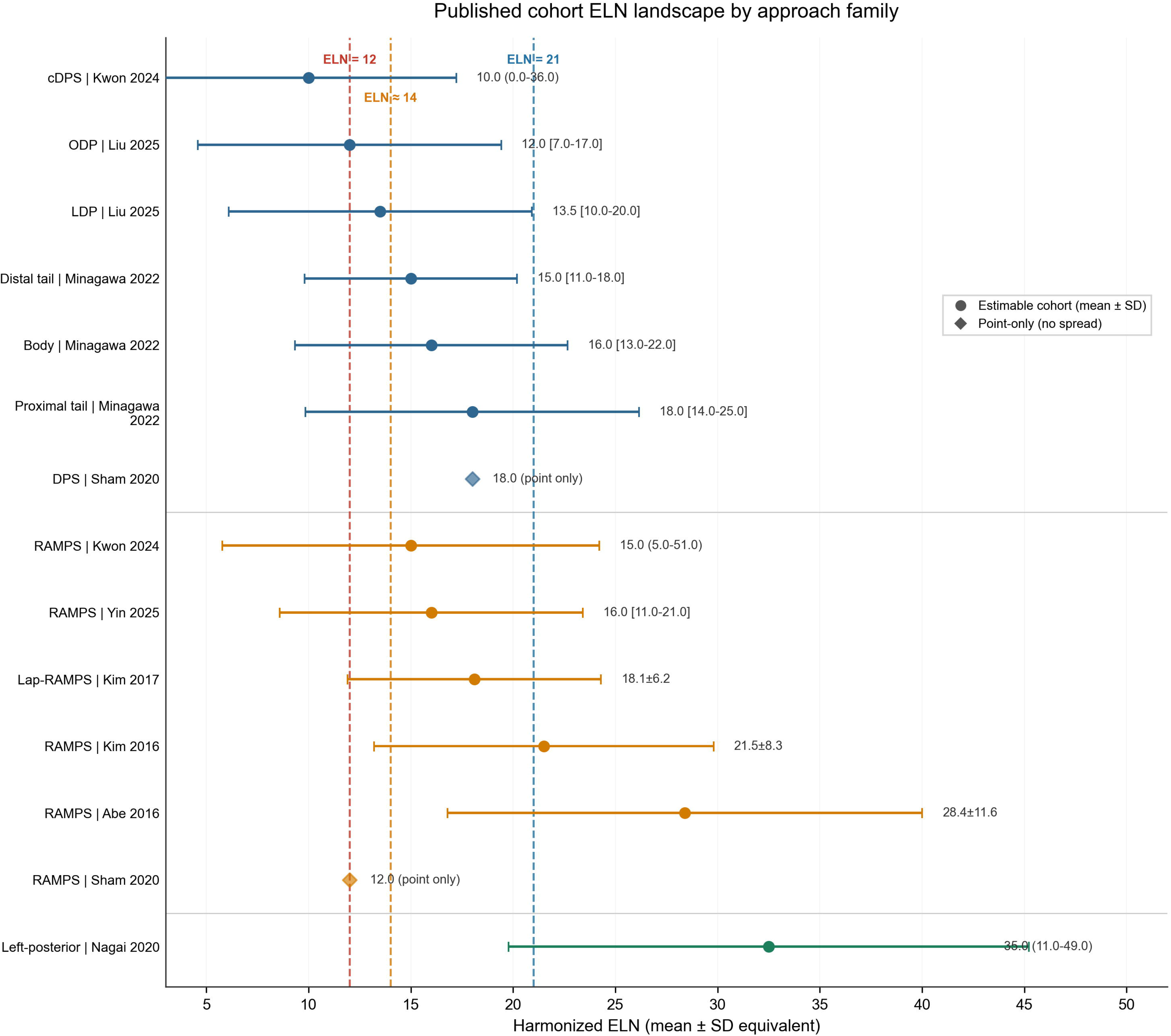
Published-cohort ELN landscape by surgical approach family. Harmonized cohort-level mean+/-SD equivalents across 17 studies for conventional distal pancreatectomy (DP), radical antegrade modular pancreatosplenectomy (RAMPS), and deep retroperitoneal/left-posterior approaches (single cohort, n=9), displayed against the SEER-derived thresholds ELN=12, 14, and 21.

### Theoretical Modeling of Surgical Accessibility

The preceding distribution-derived analysis estimated that the deep retroperitoneal/left-posterior family, derived from a single cohort (Nagai 2020, n=9), attains the SEER-derived higher-yield threshold most often (82.7%, model-estimated). We next asked whether a specific standardised retroperitoneal maneuver-the Mattox maneuver for left-sided retroperitoneal exposure-could mechanistically explain and bridge this accessibility gap.

### Anatomical accessibility and counterfactual

Modeling of station-level accessibility yielded estimated model ELN of 16 (range 12-18) for conventional DP, 21 (range 18-24) for RAMPS, and 27 (range 26-34) for the Mattox maneuver (Supplementary Figure S8A; Table 3). The additional stations accessible via Mattox (Delta-S) included #11p root, #8a, #14, #11p enhanced, #9 partial, #8p, #16b1, #16a2, and #9 full. Counterfactual removal of Delta-S reduced Mattox ELN from 27 to 12, which is below the ELN=21 threshold, confirming that deep clearance is irreplaceable rather than redundant (Supplementary Figure S8A, open symbol). Sensitivity analysis using worst-case (ELN=26, Delta-S=21) and best-case (ELN=34, Delta-S=9) bounds preserved the qualitative conclusion.

**Table 3.**
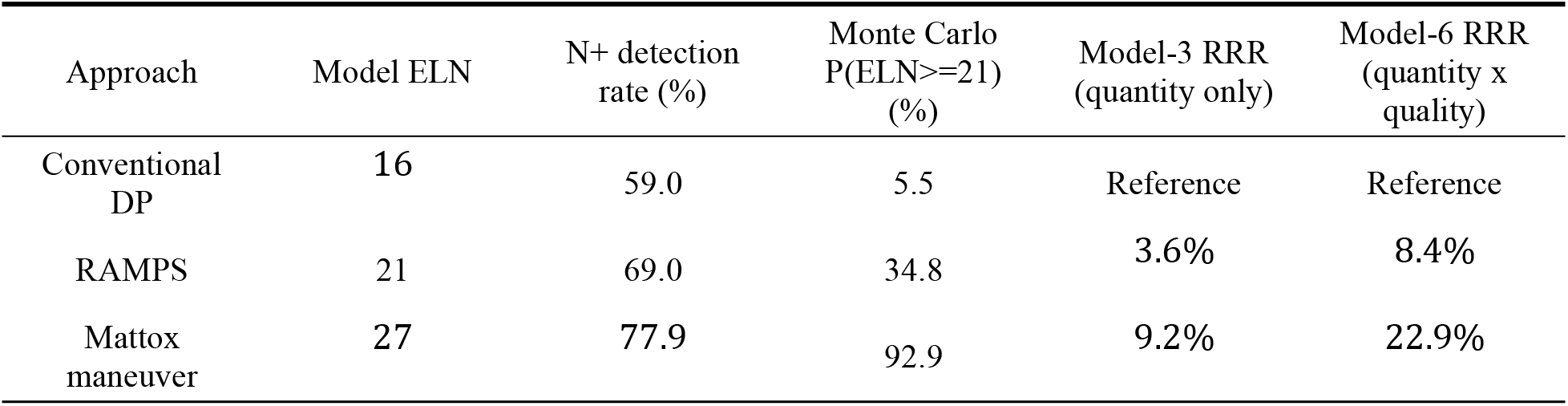
Theoretical model ELN, N+ detection rate, Monte Carlo P(ELN>=21), and relative risk reduction by approach (Mattox values are model predictions, not clinical observations).

### Dose-response and survival benefit modeling

Using the SEER-derived dose-response function, the per-node hazard reduction attenuated above the changepoint (spline-derived HR 0.993-0.995 per node above ELN 21 versus 0.992 below; Supplementary Table ST02). The Model-3 RRR (quantity-only) was 9.2% for Mattox versus DP and 3.6% for RAMPS versus DP (Supplementary Figure S8B). The dose-response was extrapolated to ELN=27 where SEER data are sparse; this extrapolation is a limitation.

The quantity-only RRR (9.2%) reflects the modeled ELN difference through the SEER dose-response curve, whereas the Model-6 RRR (22.9%) additionally incorporates a clearance-depth (HR_quality) term; the difference reflects the structural assumption that deep clearance provides an independent quality benefit, not an empirical observation.

### Staging accuracy and Monte Carlo simulation

Modeling of N+ detection probability assumed an i.i.d. positive-node probability of 0.0543 per node, calibrated to the weighted N+ rate of 54.2% at a reference ELN of 14. At ELN=16 (model DP), the N+ detection rate was 59.0%; at ELN=27 (model Mattox), it was 77.9%, representing an absolute improvement of 18.9 percentage points in N+ detection (Supplementary Figure S9A; Table 3). This staging-accuracy estimate aligns directionally with the empirical POS step described above.

Monte Carlo simulation (10 000 runs) of ELN distributions produced P(ELN>=21) of 5.5% for DP, 34.8% for RAMPS, and 92.9% for the Mattox maneuver (Supplementary Figure S9B; Table 3). The directionality is consistent with the distribution-derived attainment rates (16.5% / 40.0% / 82.7%).

### HR decomposition and the RAMPS paradox

To explain the apparent paradox that RAMPS increases ELN without improving overall survival (Kwon 2024, P=0.853), we decomposed the modeled HR into a quantity component (HR_quantity, derived from the SEER dose-response) and a clearance-depth quality component (HR_quality, reflecting deep-station coverage; Supplementary Figure S8C). RAMPS improved HR_quantity (more ELN) but had limited deep-station coverage (#9 and #14 partial or absent), yielding Model-6 RRR of 8.4% versus DP. Mattox improved both quantity and quality, yielding Model-6 RRR of 22.9% versus DP. The Kwon propensity score match study result (RAMPS ELN gain of 5.0 nodes, P=0.001; OS P=0.853) is therefore consistent with a model in which the quantity-only benefit of an anterior approach modification is below the magnitude required for a detectable survival effect, whereas a maneuver that simultaneously changes clearance depth may, in principle, cross that threshold. This interpretation is theoretical and testable: if correct, the proportion of deep-station ELN should correlate with overall survival independently of total ELN count

### Internal validation

Sensitivity analysis using one-at-a-time parameter perturbation showed that the dominant driver of RRR variability was the HR per ELN above the changepoint (RRR swing 30.0 percentage points across the plausible range 0.95-1.00), followed by the clearance-quality weight (swing 5.2 percentage points) and the HR per ELN below the changepoint (swing 2.9 percentage points) (Supplementary Figure S7A). Other parameters, including the additional station counts for Mattox (#9, #11p, #14, #16), produced RRR swings below 1.5 percentage points. Parametric bootstrap (500 replicates, 1 000 inner simulations per replicate) produced Mattox P(ELN>=21) 95% percentile CI of 91.1-94.4% and DP 95% CI of 4.9-7.8% (Supplementary Figure S7B). Cross-validation against three external evidence points (Minagawa median ELN=16 [12], Portelli meta-analytic RAMPS-ELN gain=5.91 [33], Nagai left-posterior ELN=35 [23]) showed directional concordance (Supplementary Figure S7C). These results support parameter robustness of the modeling; they do not establish external validity or clinical effectiveness.

## Discussion

This study combined SEER-based survival analysis with a structured evidence synthesis and a hypothesis-generating theoretical framework. Findings support a dual-threshold interpretation: ELN>=12 as a minimum benchmark and ∼21 as a higher-yield target (60-month RMST gain 2.60 months). The framework suggested that a standardised retroperitoneal approach (Mattox maneuver) could bridge this gap.

ELN=12 is supported by guideline recommendations [13,14] and the empirical POS step (HR 1.403 for POS>=1 versus POS=0) [34,35]. The ∼21-node target is hypothesis-generating rather than definitive: the changepoint had a wide bootstrap CI (6.0-35.0) and the spline attenuated above 21. HR 0.964 per 5 ELN supports this interpretation.

The ELN-survival association is not causal. Increasing ELN may improve staging accuracy, reduce stage migration, and serve as a surrogate for surgical quality [36], consistent with the observed survival differences. The theoretical modeling uses the empirical dose-response to explore how surgical accessibility constrains threshold attainment.

Distribution-derived (model-estimated) attainment rates (16.5%/40.0%/82.7%) and Monte Carlo P(ELN>=21) (5.5%/34.8%/92.9%) are directionally consistent. Both derive from overlapping literature inputs and are not fully independent, yet this convergence supports the hypothesis that retroperitoneal clearance enables threshold attainment.

The RAMPS paradox is central: Kwon 2024 showed that RAMPS increases ELN without improving survival [17], the pattern predicted by the quantity-only Model-3 when clearance depth is held constant. Model-6 additionally incorporates HR_quality, exceeding the magnitude for a detectable survival effect if the quality hypothesis is correct, but this remains a testable prediction.

ELN should not be equated with indiscriminate lymphadenectomy. Trials of extended versus standard lymphadenectomy for pancreatic head cancer have not shown survival benefit [37,38]. These studies addressed head tumours, not body/tail retroperitoneal depth (Supplementary Table ST05). The framework is consistent: quantity-only dissection produces little benefit; changing depth (Mattox) might differ, requiring prospective evaluation.

This study has limitations. SEER lacks neoadjuvant therapy, margin status, surgeon experience, and hospital volume data, which may confound the ELN-survival association. The evidence synthesis is limited by study-design heterogeneity: the deep retroperitoneal/left-posterior family is represented by a single small cohort (Nagai 2020, n=9) [23], so its model-estimated attainment rates are low-certainty, trend-level values; two artery-first reports (Takaori 2016 [27]; Ome 2019 [24]) provide mechanistic context only and contribute no ELN data. Mattox predictions are model-based, not clinical observations; the N+ detection model assumed i.i.d. per-node positivity whereas nodal metastases cluster. Model parameters were calibrated from literature and have not been externally validated.

ELN>=12 is a minimum quality benchmark after resection for pancreatic body/tail PDAC, and ∼21 a potential higher-yield target. Conventional DP carries an accessibility gap; theoretical modeling suggests retroperitoneal approaches such as the Mattox maneuver could bridge this gap. ELN alone is insufficient; retroperitoneal clearance depth may enable threshold attainment. These predictions are hypothesis-generating and require prospective validation.

## Supporting information

STROBE_Checklist_merged_ELN_cohort

## Data Availability

The primary data used in this study (e.g., survival and node count data) were obtained from the publicly available, de-identified Surveillance, Epidemiology, and End Results (SEER) database (accessible at https://seer.cancer.gov/) and previously published, aggregated data (e.g., Nagai et al.). These data are available from the respective sources. The quantitative model outputs and synthesis data generated in this work are available in the manuscript and its supplementary materials, or upon reasonable request to the authors.

## Declarations

### Ethics approval and consent to participate

This study used de-identified public SEER data and was exempt from institutional review board approval and individual informed consent according to the SEER data-use agreement.

### Consent for publication

Not applicable; this manuscript contains no individual patient data.

### Funding

This work was supported by the Discipline Cluster of Oncology, Wenzhou Medical University, China. The funder had no role in study design, data collection, analysis, interpretation, or manuscript preparation.

### Conflict of interest

The authors declare no competing interests.

### Data availability

The SEER data used in this study are available through the SEER Research Plus database after completion of the SEER data-use agreement. Aggregate model parameters and analysis code are available from the corresponding author upon reasonable request.

### Code availability

Analysis code is available at the corresponding GitHub repository (https://github.com/vermouth-SVG/PDAC-ELN-analysis).

### Preregistration

This study was not preregistered in an independent institutional registry.

### Reporting guideline

This study is reported in accordance with the STROBE guideline. A completed STROBE checklist is provided as a supplementary file.

### Use of artificial intelligence

AI-assisted tools were used to support code formatting, programming assistance, language editing, and manuscript formatting during study preparation. All study design decisions, data verification, statistical analyses, clinical interpretation, and manuscript content were conducted and approved by the authors, who take full responsibility for the integrity of the work.

### Author contributions

Y.Z. proposed the original clinical research question and supervised the study. X.Y. designed the analysis, performed SEER data extraction and statistical modeling, and drafted the manuscript. W.Y., J.W., Y.W., J.F., and G.M.K. contributed to study design, literature review, data interpretation, and critical revision of the manuscript. All authors reviewed and approved the final version and agree to be accountable for all aspects of the work.

## Acknowledgements

The authors thank the SEER Program registries for providing access to public cancer registry data.

## Previous communication

This work has not been presented previously.

## Supplementary materials

Supplementary Figure S1. SEER model performance: calibration curves at 12, 36, and 60 months, decision-curve analysis, and bootstrap C-index.

Supplementary Figure S2. ELN distribution stratified by stage and surgical procedure.

Supplementary Figure S3. Overlap-weighted restricted mean survival time (RMST) analysis by ELN threshold. Panel A: bar chart of RMST gain for ELN>=12, ELN>=14, and ELN>=21 across 12-, 36-, and 60-month horizons. Panel B: heatmap of weighted Delta RMST by ELN threshold and horizon.

Supplementary Figure S4. Surgical approach profile across the 17-study synthesis (forest plot of ELN means and ranges).

Supplementary Figure S5. Anchor attainment matrix mapping each study to the model-estimated ELN>=21 attainment rate.

Supplementary Figure S6. Positive lymph node (POS) analysis. Panel A: step effect of positive lymph nodes (relative hazard for POS=0 versus POS>=1). Panel B: adjusted spline curve for the POS-mortality association. Panel C: Kaplan-Meier curves by POS tier (POS=0, POS=1-2, POS=3-4, POS>=5) with number-at-risk table. Panel D: weighted Delta RMST by POS contrast relative to POS=0 across 12-, 36-, and 60-month horizons.

Supplementary Figure S7. Internal validation (theoretical model). Panel A: tornado plot of one-at-a-time parameter perturbation, showing the dominant role of HR per ELN above the changepoint (RRR swing 30.0 percentage points). Panel B: bootstrap distribution (500 replicates, 1 000 inner simulations per replicate) of Mattox P(ELN>=21) with 95% percentile CI 91.1-94.4%. Panel C: cross-validation of model predictions against external evidence points (Minagawa median ELN, Portelli meta-analytic ELN gain, Nagai left-posterior ELN).

Supplementary Figure S8. Anatomical accessibility, counterfactual, and HR decomposition (theoretical model). Panel A: model ELN estimates for DP (16), RAMPS (21), and Mattox (27), with open symbol showing the counterfactual Mattox ELN after removal of Delta-S. Panel B: SEER-derived dose-response curve. Panel C: HR decomposition into quantity and clearance-depth components.

Supplementary Figure S9. Staging accuracy and Monte Carlo simulation (theoretical model). Panel A: N+ detection probability as a function of ELN. Panel B: Monte Carlo simulation (10 000 runs) ELN distributions and P(ELN>=21).

## Notes

### Competing Interest Statement

The authors have declared no competing interest.

### Author Declarations

The Institutional Review Board (IRB) of The First Affiliated Hospital of Wenzhou Medical University waived ethical approval for this work. This study is based exclusively on the publicly available, de-identified Surveillance, Epidemiology, and End Results (SEER) database and previously published, aggregated data (e.g., Nagai et al.); no new patient-level data were collected, and no protected health information was accessed.

### Summary of Updates

We have revised the manuscript to clarify the conceptual distinction between "artery-first" and "deep retroperitoneal/left-posterior" approaches in distal pancreatectomy. Upon re-evaluation of the literature, we confirmed that two previously grouped references (Takaori 2016 and Ome 2019) describe artery-first techniques that address vascular control sequence but do not alter the retroperitoneal dissection plane, and therefore should not be classified under the deep retroperitoneal approach family. Only Nagai 2020 represents a true deep retroperitoneal/left-posterior approach with quantitative lymph node data (n=9, ELN 35). Key revisions include: 1) Updated the Abstract, Introduction, Methods, Results, and Limitations sections to use precise terminology distinguishing artery-first (conceptual and technical support) from deep retroperitoneal/left-posterior (quantitative family) approaches. 2) Added explicit disclosure that the deep retroperitoneal/left-posterior family is represented by a single small cohort (Nagai 2020, n=9), and that associated attainment rates are low-certainty, trend-level, model-based estimates. 3) Clarified in the Methods section that the two artery-first publications serve only as mechanistic support and do not contribute quantitative ELN data. 4) Updated Supplementary Tables ST03, ST04, and ST09 with corrected family names and appended footnotes declaring the artery-first references' non-quantitative role. 5) Updated Figure 2 legend to note the single-cohort basis (n=9). 6) Updated the presentation slides to reflect revised terminology and evidence caveats. No numerical values, figures, reference numbering, or core analytical results were altered in this revision.

