## Supplementary material for "Deep Fascia Plane Dissection and Mattox Maneuver for Lymphadenectomy in Pancreatic Body–Tail Cancer: SEER-Based Evidence, Anatomical Rationale, and Quantitative Modeling": STROBE_Checklist_merged_ELN_cohort

**STROBE Statement-Checklist of items that should be included in reports of cohort studies**

*Manuscript: Examined Lymph Node Threshold in Left-Sided PDAC: SEER-Based Identification with Theoretical Modeling of Surgical Accessibility*

*Note: Locations are given by manuscript section, table, figure, or supplementary item. Page and line numbers can be updated by the authors at the production-proofing stage.*

*Checklist source: STROBE checklist for cohort studies. This file is intended as a reporting checklist for submission and editorial review.*

| **Section/topic** | **Item no.** | **STROBE recommendation** | **Location in manuscript** |
| --- | --- | --- | --- |
| Title and abstract | 1(a) | Indicate the study design with a commonly used term in the title or abstract. | Title identifies a SEER-based cohort study with theoretical modeling; Abstract, Method. |
| Title and abstract | 1(b) | Provide in the abstract an informative and balanced summary of what was done and what was found. | Abstract, Introduction/Method/Results/Conclusion. |
| Introduction | 2 | Explain the scientific background and rationale for the investigation being reported. | Introduction, paragraphs 1-4. |
| Introduction | 3 | State specific objectives, including any prespecified hypotheses. | Introduction, final paragraph: SEER-based survival analysis plus hypothesis-generating theoretical modeling. |
| Methods | 4 | Present key elements of study design early in the paper. | Methods, Study design and population. |
| Methods | 5 | Describe the setting, locations, and relevant dates, including periods of recruitment, follow-up, and data collection. | Methods, Study design and population: SEER 17 registries, 2000-2022; median follow-up 70.9 months (reverse Kaplan-Meier). |
| Methods | 6(a) | Give the eligibility criteria, and the sources and methods of selection of participants. | Methods, Study design and population: aged >=18, pancreatic body/tail PDAC, surgical resection; exclusion of cases with missing/invalid survival, ELN, or POS data (POS>ELN: 0 cases). |
| Methods | 7 | Clearly define all outcomes, exposures, predictors, potential confounders, and effect modifiers. | Methods, Threshold identification and survival modelling; Surgical approach evidence synthesis. |
| Methods | 8 | For each variable of interest, give sources of data and details of methods of assessment. | Methods, Threshold identification: log-rank grid search, segmented Cox, restricted cubic spline, multivariable Cox. Supplementary Tables ST01-ST02 contain ELN-bin survival, log-rank threshold grid, exact OS curve, ELN and POS Cox models, spline HR curves, and POS-stratified RMST differences. |
| Methods | 9 | Describe any efforts to address potential sources of bias. | Methods, RMST and POS analyses: overlap weighting and IPTW; bootstrap CIs. |
| Methods | 10 | Explain how the study size was arrived at. | Methods, Study design and population: final analytic cohort n=5107 after exclusions. |
| Methods | 11 | Explain how quantitative variables were handled in the analyses. | Methods, Threshold identification: ELN treated as continuous and as binary cut-points. |
| Methods | 12(a) | Describe all statistical methods, including those used to control for confounding. | Methods: multivariable Cox, OW/IPTW RMST, segmented Cox changepoint, RCS, POS step+spline, Monte Carlo, sensitivity/bootstrap/cross-validation. |
| Methods | 12(b) | Describe any methods used to examine subgroups and interactions. | Methods: stratified ELN by stage and procedure (Supplementary Figure S2); POS-stratified analyses (Supplementary Table ST02). |
| Methods | 12(c) | Explain how missing data were addressed. | Methods: SEER data completeness; cases with missing/invalid survival, ELN, or POS data excluded (n=88; no cases excluded for POS/ELN>1). |
| Methods | 12(d) | If applicable, describe loss to follow-up and sensitivity analyses. | Methods: bootstrap CIs; sensitivity analysis one-at-a-time parameter perturbation. |
| Methods | 12(e) | Describe any methods used to assess robustness of the analyses (e.g., sensitivity analyses). | Methods, Theoretical modeling framework: sensitivity, bootstrap, cross-validation; supplementary Table ST08 (Mattox model). |
| Results | 13(a) | Report numbers of individuals at each stage of the study. | Results, Cohort characteristics: n=5107 analytic cohort. |
| Results | 14(a) | Give characteristics of study participants and information on exposures and potential confounders. | Table 1; Results, Cohort characteristics. |
| Results | 14(b) | Indicate number of participants with missing data for each variable of interest. | Table 1 footnotes; cohort exclusions stated in Methods. |
| Results | 15 | Report numbers of outcome events or summary measures over time. | Results, Cohort characteristics: 3630 deaths (71.1%); survival modelling results. |
| Results | 16(a) | Give unadjusted estimates and, if applicable, confounder-adjusted estimates and their precision. | Results, ELN thresholds: HR 0.964 (95% CI 0.949-0.980); Table 2. |
| Results | 16(b) | Report category boundaries when continuous variables were categorized. | Results, ELN thresholds: ELN=12 (binary), ELN=21 (changepoint); ELN bins in Supplementary Figure S2. |
| Results | 16(c) | If relevant, consider translating estimates of relative risk into absolute risk for a meaningful time period. | Results, RMST: 60-month delta-RMST 2.60 months for ELN>=21. |
| Results | 17 | Report other analyses done (e.g., analyses of subgroups and interactions, and sensitivity analyses). | Results, POS analysis (Supplementary Table ST02); Theoretical Modeling of Surgical Accessibility (Supplementary Tables ST03-ST07 evidence synthesis; Table ST08 Mattox model); Internal validation (Supplementary Figure S7). |
| Discussion | 18 | Give a cautious overall interpretation of results considering objectives, limitations, multiplicity of analyses, results from similar studies, and other relevant evidence. | Discussion, paragraphs 1-7; limitations explicitly enumerated in paragraph 6. |
| Discussion | 19 | Discuss implications of the results for future research and clinical practice. | Discussion, paragraph 7 (clinical implications); Conclusion. |
| Other | 20 | Give the source of funding and the role of the funders; present conflicts of interest. | Declarations: Funding; Conflict of interest. |
| Other | 21 | Give the ethical approval status and informed consent procedures. | Declarations: Ethics approval and consent to participate. |
| Other | 22 | Describe the role of the study sponsors and the use of artificial intelligence tools. | Declarations: Use of artificial intelligence; Author contributions. |
